# White matter microstructure mediates the link between rest-activity rhythm instability and mania symptoms in individuals at risk for bipolar disorder

**DOI:** 10.64898/2026.01.13.26343956

**Authors:** João Paulo Lima Santos, Lara Labardini, Lauren Keller, Simey Chan, Danella Hafeman, Brant P Hasler, Meredith Wallace, Adriane M. Soehner

## Abstract

**Background:** Adolescence and young adulthood are critical periods for the emergence of bipolar disorder (BD). Instability in sleep and rest-activity rhythms is associated with elevated risk for BD, yet little is known about the neural correlates of this vulnerability. White matter organization in fronto-limbic pathways, which support mood regulation and show sensitivity to sleep/rest-activity rhythm disruption, offers a promising avenue for investigation.

**Methods:** Participants (16-24y;N=112) recruited across a spectrum of mania vulnerability (MOODS-SRL) completed 14 days of actigraphy followed by a neuroimaging assessment. Diffusion MRI was used to derive Neurite Orientation Dispersion and Density Imaging, focusing on the Orientation Dispersion Index (ODI) of the cingulum bundle, uncinate fasciculus, and forceps minor. Clinician-rated symptoms of mania and depression were assessed at baseline and 6-months follow-up. We evaluated whether the links between actigraphy-derived sleep duration variability, sleep onset variability, and Circadian Function Index (CFI; index of circadian rhythm robustness) and white matter ODI were moderated by baseline mania vulnerability, and tested whether baseline ODI predicted 6-month mood symptoms.

**Results:** At higher levels of mania vulnerability, lower CFI (greater rest-activity instability) associated with higher ODI (greater white matter disorganization) in the cingulum bundle (β=-0.22;P=0.029) and uncinate fasciculus (β=-0.22;P=0.020). Moreover, higher uncinate fasciculus ODI predicted greater mania symptoms at 6-month follow-up and fully mediated the CFI-mania symptom association (β=-0.10;95%CI[-0.14,-0.02]).

**Conclusions:** Rest-activity instability may disrupt fronto-limbic white matter, increasing risk to mania in those at elevated vulnerability for BD. Stabilizing rest-activity rhythms in at-risk individuals may help preserve white matter integrity and mitigate BD risk.

## 1. INTRODUCTION

Bipolar disorder (BD) is a highly impactful and persistent mental illness that often emerges during adolescence and early adulthood(Grande et al., 2016; Sajatovic, 2005). Key risk factors include a family history of BD and a lifetime history of subthreshold mood symptoms(Axelson et al., 2015; Birmaher et al., 2018; Grande et al., 2016; Vieta et al., 2018), the latter reflecting an enduring vulnerability that contributes to psychological distress and impairment that can persist for years before meeting full diagnostic criteria for BD(Faedda et al., 2015; SAMHSA, 2007; Skjelstad et al., 2010). In addition, BD is associated with several adverse outcomes, including elevated suicide risk and long-term functional difficulties that may persist even during remission(Fagiolini et al., 2005; McIntyre & Calabrese, 2019; Zhong et al., 2024). Therefore, identifying modifiable markers of vulnerability is critical to inform prevention and early intervention efforts, which could potentially delay or mitigate long-term adverse outcomes. In this context, growing evidence highlights instability in sleep and rest-activity rhythms as additional, dynamic, and modifiable vulnerability markers for BD risk(Lima Santos et al., 2025; Tonon et al., 2024).

Sleep and rest-activity rhythms fluctuate in response to environmental demands, behavior, and internal factors, producing dynamic instability(Aguiar et al., 2021; Melo et al., 2016; Ng et al., 2015; Ortiz et al., 2025). In BD, such instability has been shown to occur during acute mood states, persist between episodes, and precede recurrence(Melo et al., 2016; Ng et al., 2015; Ortiz et al., 2025), serving as a behavioral marker of underlying biological vulnerability. In addition, our recent work(Lima Santos et al., 2025) showed that the association between sleep/rest-activity rhythms instability and mood symptoms may be amplified in individuals with underlying, persistent vulnerabilities, with stronger associations observed in adolescents with a family history of BD relative to those without. This heightened vulnerability is particularly important because it suggests that disruptions in sleep and rest-activity rhythms, which are common in adolescents and young adults, may compound risk in those already showing early vulnerability to BD. Importantly, these relationships also have potential implications for the neurobiology of BD(Harrison et al., 2018), as sleep and rest-activity rhythms are critical for brain structure and function(Reyes et al., 2022; Shen et al., 2023; Telzer et al., 2015). Factors that amplify the negative effects of sleep and rest-activity rhythms instability on the brain may further disrupt neural circuits supporting mood and emotional regulation - processes central to BD - and thereby increase susceptibility to its onset. However, it remains unclear whether the influence of sleep and rest-activity rhythms on the brain differs across levels of BD risk.

The brain plays a central role in the pathology of BD, with converging evidence from neuroimaging, neuropathological, and lesion studies highlighting widespread structural and functional abnormalities across fronto-limbic networks(Drevets et al., 2008; Goldstein & Walker, 2014; Harrison et al., 2018; Ng et al., 2009; Tkachev et al., 2003). Among these findings, brain white matter abnormalities have been among the most consistent, reflecting disrupted connectivity between regions involved in emotional regulation, reward processing, and executive control. Reductions in fractional anisotropy, a conventional diffusion tensor imaging related to overall fiber collinearity(Jones & Leemans, 2010) that has shown negative relationships with severity of symptoms, are frequently observed in tracts such as the cingulum bundle(Lima Santos et al., 2022; Wang et al., 2008), corpus callosum(Videtta et al., 2023), and uncinate fasciculus(Foley et al., 2018) which are key pathways integrating prefrontal and limbic regions. Alterations in the integrity of these tracts also precede BD recurrence, suggesting they may serve as early indicators of vulnerability(Lima Santos et al., 2022; Santos et al., 2022). Notably, studies have also shown a strong relationship between sleep, rest-activity rhythms, and white matter(Callow et al., 2025; Guldner et al., 2023; Jamieson et al., 2020; McMahon et al., 2021; Reyes et al., 2022; Telzer et al., 2015) . Yet, these relationships remain largely unexplored in the context of sleep and rest-activity rhythms instability’s unique associations in BD risk(Lima Santos et al., 2025). Examining how sleep and rest-activity rhythms instability relate to the microstructure of fronto-limbic white matter pathways may therefore provide the neural substrates necessary to understand how these processes compound vulnerability to BD.

The aims of this study were twofold. First, we examined whether the association between sleep and rest-activity rhythms instability (sleep duration, sleep onset, and circadian robustness) and white matter microstructure in key emotion regulation tracts implicated in BD (cingulum bundle, forceps minor, and uncinate fasciculus) varied across a spectrum of lifetime mania vulnerability as measured by the Mood Spectrum Self-Report Lifetime Mania Scale (MOODS-SRL; Dell’Osso et al., 2002). Second, we tested whether alterations in the white matter tracts vulnerable to sleep and rest-activity rhythms instability predicted greater depressive and/or mania symptoms at a 6-month follow-up and mediated the association between sleep/rest–activity instability and subsequent mood symptoms. To address these aims, we used Neurite Orientation Dispersion and Density Imaging (NODDI; Timmers et al., 2016; Zhang et al., 2012), an advanced diffusion MRI technique that provides more specific measures of white matter microstructure than conventional diffusion tensor imaging (FA). NODDI quantifies the structural organization of neurites (axons and dendrites) via the Neurite Density Index (NDI), reflecting axonal packing, and the Orientation Dispersion Index (ODI), reflecting the coherence of neurite orientation. In our prior work, sleep disruption was particularly linked to higher ODI (greater dispersion, lower coherence) which in turn was associated with greater mood symptoms in adolescent with no history of psychiatric disorders(Santos et al., 2025). Therefore, we hypothesized that greater sleep and rest-activity rhythms instability would be associated with higher ODI, especially in individuals with higher MOODS-SRL scores, and that higher ODI would predict increased depressive and mania symptoms six months later. We also explored associations with other sleep measures (lights out and get up time) and with other brain white matter metrics (NDI and FA).

## 2. METHODS

### 2.1. Study and participants

This study obtained approval from the Office of Research Protections at the University of Pittsburgh. The analytical sample consisted of adolescents and young adults aged 16-24 years who exhibited a range of lifetime mania vulnerability based on MOODS-SRL mania subscale, but no current or lifetime bipolar or psychosis spectrum disorder. Stratification used the following MOODS-SRL mania subscale ranges: low=0-5, medium=6-14; or high≥15. Exclusion criteria are described in the Supplemental Methods.

In this ongoing study, we used a data cut as of September/2025. A total of 114 participants completed the baseline assessment (see 2.4 Procedures). Two participants were excluded due to failing neuroimaging data quality control, leaving 112 participants in the final baseline sample. Distribution of MOODS-SRL in this sample is shown in Supplemental Figure 1. A subsample of 82 participants completed the 6-month follow-up.

### 2.2. Measures

#### 2.2.1. Lifetime mania vulnerability

Lifetime presence of mania vulnerability was assessed with the MOODS-SRL mania subscale (Dell’Osso et al., 2002). This subscale characterizes the full spectrum of mania disturbance (subsyndromal, prodromal, and syndromal) across 62 items evaluating lifetime manifestations of mania-relevant symptoms (mood elevation, increased activity and energy, irritability), behaviors, and personality traits. Items are scored as Yes (1) or No (0), indicating whether a statement had occurred during the respondent’s lifetime. Total scores were calculated for each participant.

#### 2.2.2. Psychiatric and Sleep Disorders

DSM-5 psychiatric disorders were assessed using the Kiddie Schedule for Affective Disorders and Schizophrenia(KSADS; Ambrosini, 2000) for participants ≤18years and the SCID(First, 2014) for participants >18 years.

#### 2.2.3. Mania and depression severity

Past-week symptoms of mania and depression were assessed using the Hamilton Rating Scale for Depression(HRSD; HAMILTON, 1960) and the Young Mania Rating Scale (YMRS; Young et al., 1978). Unlike the MOODS-SRL, which captures lifetime vulnerability and trait-like mania spectrum features, the YMRS assesses current, state-level severity of mania symptoms and is commonly used to diagnose and monitor acute mania and treatment response.

#### 2.2.4. Sleep diary

Daily self-reports captured standard data elements recommended by the consensus sleep diary(Carney et al., 2012) (e.g., bedtime, lights off time, sleep offset time, get up time) for each bout of sleep. Participants were asked to mark each bout of sleep as their main sleep internal or naps. Diaries were distributed via text each morning at their preferred time based on their habitual weekend and weekday rise times. Sleep diaries had a 12-hour response window.

#### 2.2.5. Actigraphy

A wrist accelerometer (Geneactiv Original; ActivInsights, England) was worn continuously on the non-dominant wrist. The watch can detect activity, skin temperature, and environmental white light. Sampling frequency was 50 hz. Participants were instructed to wear the device continuously except when fully submerged in water or during activities that might damage the watch, such as contact sports. Participants were instructed to indicate lights off time (beginning of sleep attempt) and get up time (physically getting out of bed) by pressing the button on the top of the device (event marker). Tracking days (noon-noon) were omitted if they contained ≥4 hours of non-wear during active intervals and ≥15 minutes during the sleep interval(Patel et al., 2015).

#### 2.2.6. Sleep estimation

Accelerometer data were processed in R using GGIR v3.1.2(van Hees et al., 2025) and a custom, semi-automated rest interval detection pipeline (actiSleep). First, GGIR generated initial estimates of sleep intervals based on activity (Cole-Kripke algorithm), identified non-wear periods, and generated an activity, light, and temperature time series down-sampled to 1-minute epochs with imputed activity for non-wear/invalid periods. Next, actiSleep implemented a hierarchical system rest interval determination based on a prior standardized scoring algorithm(Patel et al., 2015). Multiple indicators of rest onset and offset times (event marker press, sleep diary, changes in white light, and the Cole-Kripke algorithm) were aligned, then compared for agreement and weighted in order of priority (marker>diary>light>Cole-Kripke) to determine the rest interval onset and offset times. Actograms were generated using the new rest interval onset and offset times, and tracking days with atypical rest interval patterns were flagged for manual review. Tracking days were flagged based on short sleep bouts (<3 hours), long sleep bouts (>10 hours), missed marker presses (instances in which a marker press existed but was too far from other rest interval indicators for alignment or when onset/offset lacked any marker), and days with no detected sleep. Flagged days were edited using the same hierarchical scoring rules noted above, as needed. Lastly, GGIR was re-run using a curated sleep log containing the actiSleep-derived rest-interval start and end times and a data-cleaning file identifying excluded days, yielding the final sleep and rest–activity rhythm metrics.

The average and standard deviation (variability) of standard sleep variables were extracted: lights off time, sleep latency (minutes between lights off and sleep onset), sleep onset time, minutes awake after sleep onset, sleep offset time, get up time, snooze time (minutes between sleep offset and get up time), time in bed (time between lights off and get up time), total sleep time (time in bed minus sleep latency, wake after sleep onset, and snooze time). Primary measures were the within-subject standard deviation in sleep duration (total sleep time) and sleep onset time capture daily variability in sleep duration and timing.

#### 2.2.7. Rest-activity rhythms

Non-parametric rest-activity rhythms outcomes were derived with the nParACT R package(Smagula et al., 2018). Non-wear segments were imputed using GGIR data. Our primary outcome of interest was the Circadian Function Index (CFI; Ortiz-Tudela et al.), a composite measure of circadian activity rhythm robustness. The CFI is based on three components: 1) Intradaily Variability (IV), which reflects how much activity changes from hour-to-hour relative to overall activity variability; higher values indicate more fragmented rest-activity rhythms; 2) Interdaily Stability (IS), which reflects consistency of activity patterns from day-to-day; higher values indicate greater rest-activity rhythms stability; and 3) Relative Amplitude (RA), which reflects the average activity ratio of the active 10 hours relative to the least active 5 hours of the day; higher values indicate greater rest-activity rhythms amplitude. The CFI is calculated as the average of IS, RA, and a transformed IV component (1 - IV/2) and ranges from 0 (absence of circadian rhythmicity) to 1 (highly robust circadian rhythm).

### 2.3. Neuroimaging

#### 2.3.1. Protocol, preprocessing, and quality control

Diffusion Magnetic Resonance Imaging (MRI) data were collected using multiband Echo-planar imaging (EPI) with slice acceleration factor 4. It included 199 diffusion directions (TR=3230ms, TE=89.20ms, flip angle=78, FOV=210×210mm^2^, spatial resolution=1.5mm isotropic) with two b-values (93 with b=1500s/mm2, and 92 with b=3000s/mm2) along with 14 b=0 frames. Diffusion MRI was corrected for eddy current, subject motion, and echo-planar imaging distortion(Andersson et al., 2003; Casey et al., 2018; Hagler Jr et al., 2019; Smith et al., 2004). Quality control procedures included assessing raw data integrity (e.g., missing anatomical structures due to improper acquisition cuts), motion artifacts, susceptibility distortions, and signal dropouts, as well as reviewing post-processing outputs (e.g., tissue segmentation, skull stripping, metric maps) and inspecting the overall continuity of tracts.

#### 2.3.2. White matter tract segmentation

TractSeg(Wasserthal et al., 2018a; Wasserthal et al., 2018b) was used to reconstruct our white matter tracts of interest (cingulum bundle(Jones et al., 2013), forceps minor(Fabri et al., 2014), and uncinate fasciculus(Von Der Heide et al., 2013)) and additional tracts used for exploratory analyses (see Supplemental Material). In addition, for exploratory analyses, a tract-profile(Lima Santos et al., 2021; Yeatman et al., 2012) was derived to examine variations in fiber characteristics along each tract. Each tract profile consisted of five consecutive nodes, with each node representing 20% of the entire tract, thereby enabling the identification of specific tract segments involved in the observed relationships.

#### 2.3.3. NODDI and diffusion metrics

After preprocessing, the NODDI model was fitted using the NODDI toolbox(Fukutomi et al., 2018; Zhang et al., 2012). Orientation Dispersion Index (ODI) and Neurite Density Index (NDI) were derived for each participant. In addition, FA maps were derived for each participant using Mrtrix3(Tournier et al., 2019). For each segmented tract, the overall mean and nodal ODI, NDI, and FA were extracted. Finally, given that laterality was not part of our hypotheses, we averaged diffusion metrics across hemispheres.

### 2.4. Procedures

Participants were recruited through community advertisements, local clinics, and a participant registry. After completing preliminary screening procedures, participants (and a parent or guardian if under 18 years of age) completed informed consent/assent procedures and an eligibility evaluation that included clinician rated psychiatric diagnostic interviews (SCID or KSADS), family history, and self-report questionnaires. Eligible participants were provided with an actigraph for at-home baseline sleep monitoring over a 14-day period and completed sleep diaries. At the end of this monitoring period, neuroimaging data were collected, along with assessments of past-week mania and depression severity (YMRS and HRSD). Mania and depression assessments, two weeks of sleep tracking, and sleep diaries were repeated at the 6-month follow-up.

### 2.5. Statistical approach

To test our hypotheses, we employed linear regression models in R. Significant moderation effects (P<0.05) were further explored using the Johnson-Neyman technique(Curran et al., 2006; Johnson & Fay, 1950; Preacher et al., 2006). Mediation analyses were conducted using the PROCESS macro with bootstrapping (10,000 resamples; Hayes, 2017). To account for multiple comparisons, the False Discovery Rate (FDR) correction(Benjamini & Hochberg, 1995) was applied across all models for each outcome separately.

#### 2.5.1. Moderated associations between sleep/rest-activity rhythms instability and ODI

We tested whether mania vulnerability (mania subscale of MOODS-SRL) moderated the association between sleep or rest-activity rhythms instability (sleep duration variability, sleep onset variability, and CFI) and ODI of white matter tracts of interest (cingulum bundle, forceps minor, and uncinate fasciculus) at baseline. Models controlled for age, sex at birth, ethnicity, race, history of psychiatric disorders, family history of BD, and actigraphy variables (total tracking days and weekday/weekend ratio).

#### 2.5.2. Baseline ODI predicting follow-up mood symptoms

Models examined whether baseline ODI metrics of tracts of interest (cingulum bundle, forceps minor, and uncinate fasciculus) predicted follow-up depressive (HRSD) and mania (YMRS) symptoms six months later. Because the distributions of depressive and mania symptoms at follow-up were non-normal, scores were log-transformed before analysis. Models controlled for age, sex at birth, ethnicity, race, history of psychiatric disorders, family history of BD, and baseline mania vulnerability (MOODS-SRL). In addition, baseline depressive or mania symptoms were included as covariates in the respective models.

#### 2.5.3. Mediation analyses

Based on the significant associations (P<0.05) observed in sections *2.5.1* and *2.5.2*, mediation models examined whether ODI mediated the relationship between baseline sleep or rest-activity rhythms instability and depressive and/or mania symptoms six months later.

#### 2.5.4. Exploratory analyses

##### CFI components

To better understand which components of CFI possibly contributed to significant moderation effects in 2.5.1 models, we conducted follow-up models examining IV, IS, and RA separately.

##### Tract-profile analyses

Tract-profile analyses examined whether the significant associations observed in section *2.5.1* were focal (e.g., restricted to a few nodes) or widespread (across all nodes).

##### Other sleep and rest-activity rhythms metrics

Models analogous to those in section *2.5.1* tested whether mania vulnerability moderated the associations between other sleep or rest-activity rhythms metrics (sleep duration average, sleep onset average, lights-off time, and get-up time) and ODI of white matter tracts of interest. Significant associations (P<0.05) were further examined as potential mediators of the relationships between baseline sleep or rest-activity rhythms metrics and mania and depressive symptoms six months later, using models described in section *2.5.3*.

##### Other diffusion metrics and white matter tracts

Using models similar to Section *2.5.1*, we explored whether mania vulnerability moderated the relationship between sleep or rest-activity rhythms instability and other NODDI metrics (NDI) or FA. We also explored moderated effects involving additional white matter tracts not directly related to our primary hypotheses.

## 3. RESULTS

### 3.1. Sample characteristics

Clinical and demographic characteristics of the sample are presented in Table 1. Participants had a mean age of 21.69 years (SD=2.10) and were 58.9% female. MOODS-SR-L mania subscale scores ranged from 0 to 42, with a mean of 12.29 (SD=9.52). There were no significant demographic or clinical differences between the full baseline sample and the follow-up subsample (Table 1).

**Table 1.**
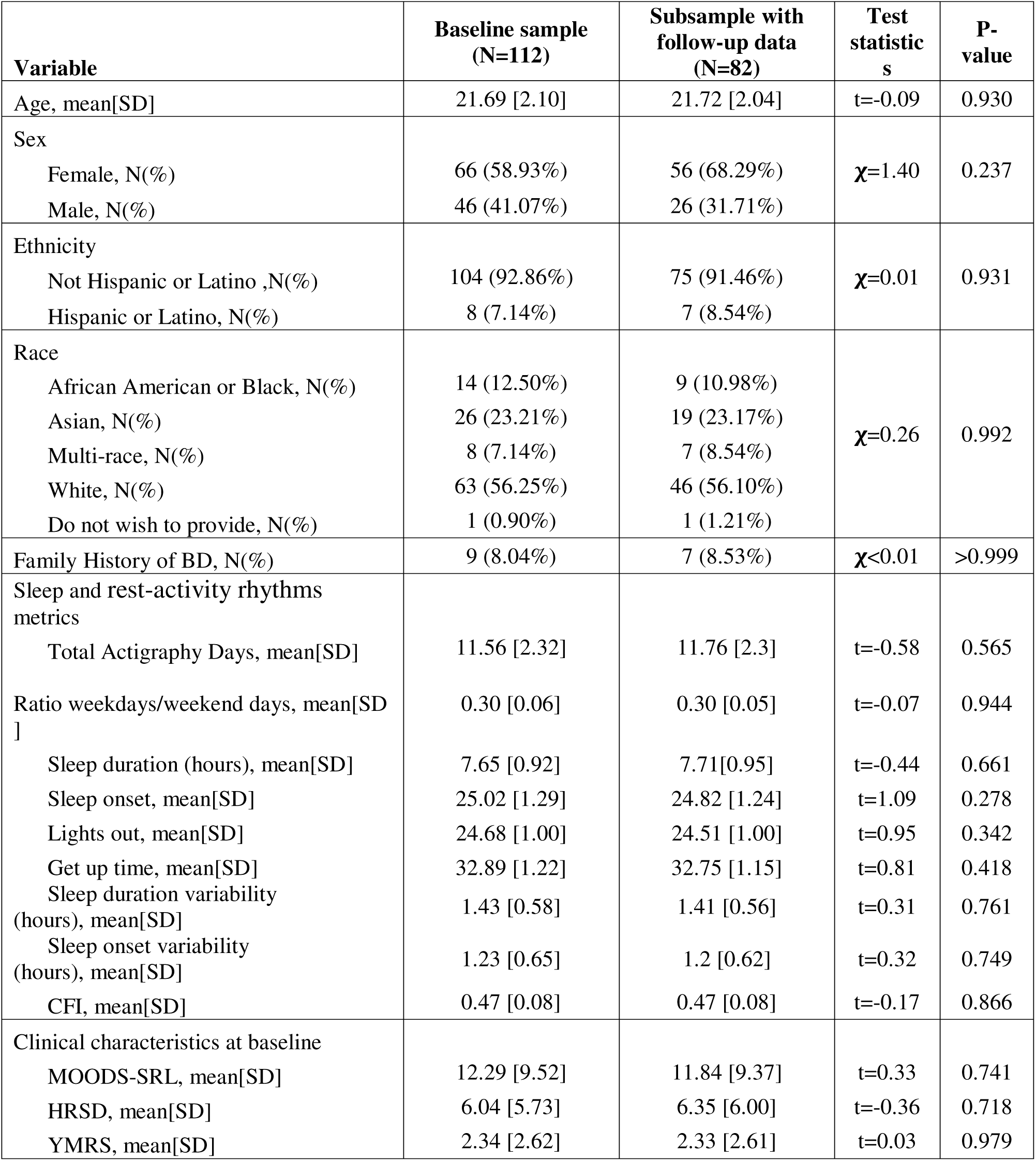
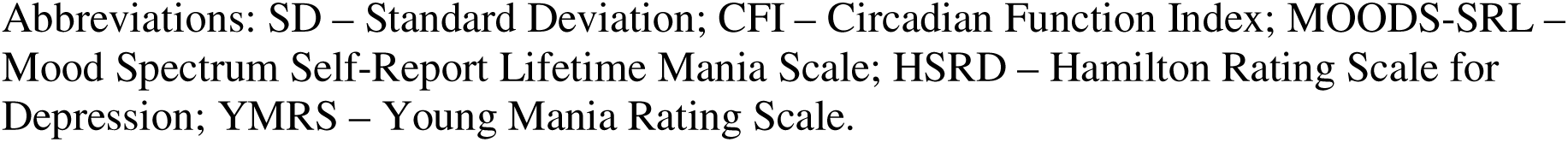
Sample characteristics and comparison between baseline sample (N=112) and subsample with follow-up data (N=82).

| Variable | Baseline sample<br>(N=112) | Subsample with<br>follow-up data<br>(N=82) | Test<br>statistic<br>s | P-value |
| --- | --- | --- | --- | --- |
| Age, mean[SD] | 21.69 [2.10] | 21.72 [2.04] | t=-0.09 | 0.930 |
| Sex |  |  |  |  |
| Female, N(%) | 66 (58.93%) | 56 (68.29%) | $\chi=1.40$ | 0.237 |
| Male, N(%) | 46 (41.07%) | 26 (31.71%) |  |  |
| Ethnicity |  |  |  |  |
| Not Hispanic or Latino ,N(%) | 104 (92.86%) | 75 (91.46%) | $\chi=0.01$ | 0.931 |
| Hispanic or Latino, N(%) | 8 (7.14%) | 7 (8.54%) |  |  |
| Race |  |  |  |  |
| African American or Black, N(%) | 14 (12.50%) | 9 (10.98%) | $\chi=0.26$ | 0.992 |
| Asian, N(%) | 26 (23.21%) | 19 (23.17%) |  |  |
| Multi-race, N(%) | 8 (7.14%) | 7 (8.54%) |  |  |
| White, N(%) | 63 (56.25%) | 46 (56.10%) |  |  |
| Do not wish to provide, N(%) | 1 (0.90%) | 1 (1.21%) |  |  |
| Family History of BD, N(%) | 9 (8.04%) | 7 (8.53%) | $\chi<0.01$ | >0.999 |
| Sleep and rest-activity rhythms<br>metrics |  |  |  |  |
| Total Actigraphy Days, mean[SD] | 11.56 [2.32] | 11.76 [2.3] | t=-0.58 | 0.565 |
| Ratio weekdays/weekend days, mean[SD]<br>] | 0.30 [0.06] | 0.30 [0.05] | t=-0.07 | 0.944 |
| Sleep duration (hours), mean[SD] | 7.65 [0.92] | 7.71[0.95] | t=-0.44 | 0.661 |
| Sleep onset, mean[SD] | 25.02 [1.29] | 24.82 [1.24] | t=1.09 | 0.278 |
| Lights out, mean[SD] | 24.68 [1.00] | 24.51 [1.00] | t=0.95 | 0.342 |
| Get up time, mean[SD] | 32.89 [1.22] | 32.75 [1.15] | t=0.81 | 0.418 |
| Sleep duration variability<br>(hours), mean[SD] | 1.43 [0.58] | 1.41 [0.56] | t=0.31 | 0.761 |
| Sleep onset variability<br>(hours), mean[SD] | 1.23 [0.65] | 1.2 [0.62] | t=0.32 | 0.749 |
| CFI, mean[SD] | 0.47 [0.08] | 0.47 [0.08] | t=-0.17 | 0.866 |
| Clinical characteristics at baseline |  |  |  |  |
| MOODS-SRL, mean[SD] | 12.29 [9.52] | 11.84 [9.37] | t=0.33 | 0.741 |
| HRSD, mean[SD] | 6.04 [5.73] | 6.35 [6.00] | t=-0.36 | 0.718 |
| YMRS, mean[SD] | 2.34 [2.62] | 2.33 [2.61] | t=0.03 | 0.979 |

Abbreviations: SD – Standard Deviation; CFI – Circadian Function Index; MOODS-SRL – Mood Spectrum Self-Report Lifetime Mania Scale; HSRD – Hamilton Rating Scale for Depression; YMRS – Young Mania Rating Scale.

### 3.2. Moderated associations between sleep/rest-activity rhythms instability and white matter coherence (ODI)

The association between CFI and ODI metrics of the cingulum bundle and uncinate fasciculus was significantly moderated by mania vulnerability (MOODS-SRL; Table 2). Johnson-Neyman analyses indicated that lower circadian rhythm robustness (lower CFI) was associated with less coherent neurite orientation (higher ODI) in both tracts, with significant effects observed for mania vulnerability scores above 20 for the cingulum bundle (Figure 1-Panel C) and above 17 for the uncinate fasciculus (Figure 1-Panel F). Trend-level FDR-corrected associations were observed between CFI and ODI of the forceps minor (Table 2). There were no associations involving sleep duration variability or sleep onset variability (Table 2).

**Figure 1.**
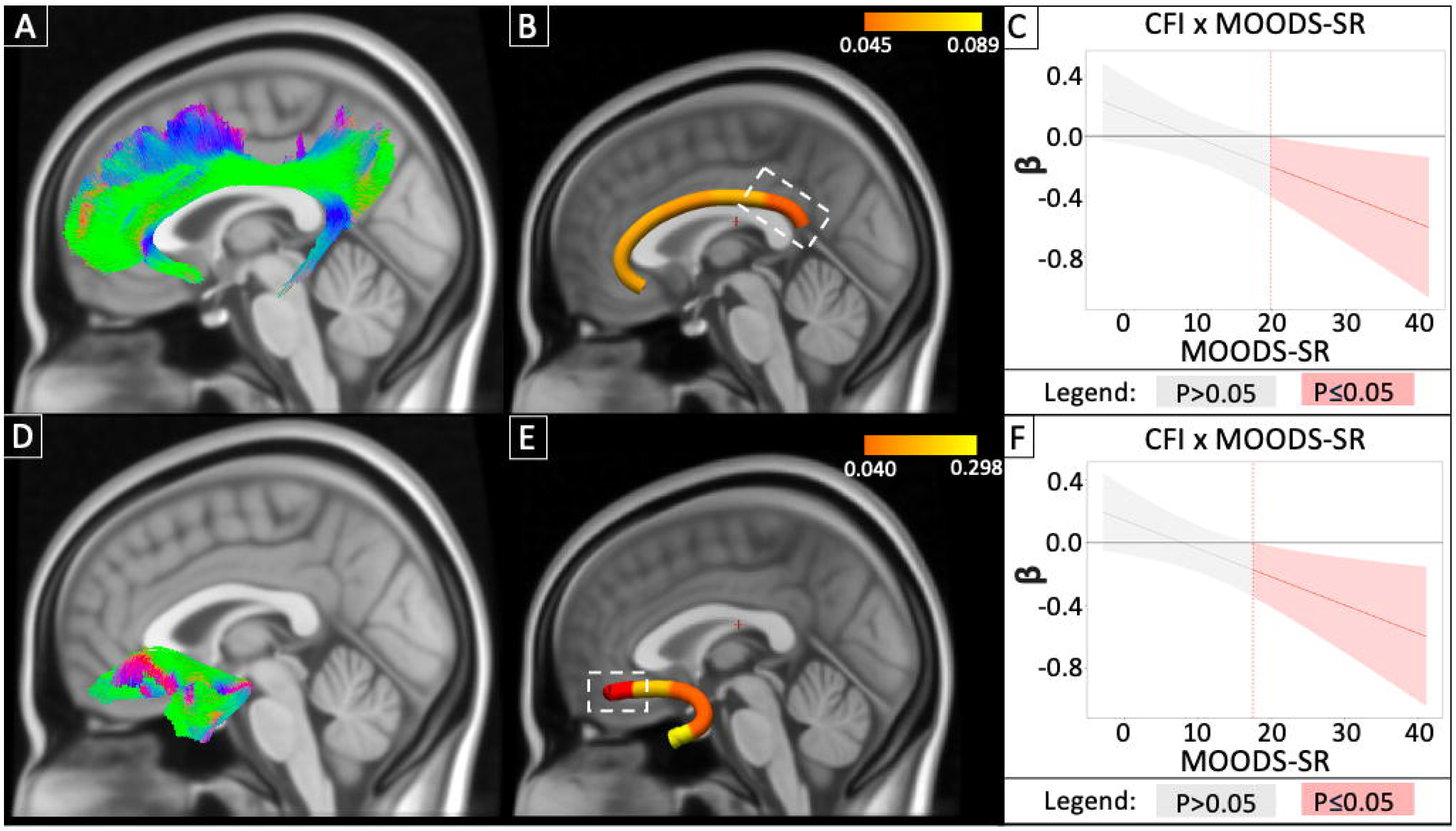
Moderated association between CFI and ODI of the cingulum bundle and uncinate fasciculus. Panels A-C show findings for the cingulum bundle, and panels D-F show findings for the uncinate fasciculus. Panels A and D display the reconstructed tracts overlaid on the standard MNI-152 1 mm brain template. Colors indicate streamline orientation: green (anterior-posterior), blue (superior-inferior), and red (right-left). Panels B and E present tract-profile effects, with the red-yellow color bar representing Q-value ranges; dotted white lines mark tract segments with Q-values<0.05. Panels C and F show Johnson-Neyman plots depicting the conditional effects of Circadian Function Index (CFI) on Orientation Dispersion Index (ODI) across levels of mania vulnerability (MOODS-SRL). The x-axis represents MOODS-SRL scores, and the y-axis represents the estimated regression coefficient for the CFI-ODI association. The solid line indicates the conditional effect, the shaded area the 95% confidence interval, and vertical dashed lines the region of significance (P<0.05).

**Table 2.** Moderated association between sleep or rest-activity rhythms instability and ODI of white matter tracts involved in emotion regulation.

| Independent Variable | Outcome | $\beta$ | P<sup>a</sup> | Q<sup>a,b</sup> | $\eta^2$<sup>c</sup> |
| --- | --- | --- | --- | --- | --- |
| Sleep duration variability | Cingulum Bundle ODI | 0.09 | 0.394 | 0.808 | 0.01 |
|  | Forceps Minor ODI | -0.06 | 0.563 | 0.808 | <0.01 |
|  | Uncinate Fasciculus ODI | 0.03 | 0.808 | 0.808 | <0.01 |
| Sleep onset variability | Cingulum Bundle ODI | 0.08 | 0.430 | 0.645 | 0.01 |
|  | Forceps Minor ODI | -0.08 | 0.414 | 0.645 | 0.01 |
|  | Uncinate Fasciculus ODI | -0.01 | 0.899 | 0.899 | <0.01 |
| CFI | Cingulum Bundle ODI | -0.22 | **0.029** | **0.044** | 0.05 |
|  | Forceps Minor ODI | -0.18 | *0.053* | *0.053* | 0.04 |
|  | Uncinate Fasciculus ODI | -0.22 | **0.020** | **0.044** | 0.06 |

Abbreviations: ODI – Orientation Dispersion Index; CFI – Circadian Function Index.
<sup>a</sup> P-values $\leq 0.05$ are reported in bold characters; P-values between 0.05 and 0.10 are reported in italics.
<sup>b</sup> Q-values indicate P-values after False Discovery Rate (FDR) correction for multiple comparisons. FDR correction was applied separately for each independent variable across all outcomes it was tested on.
<sup>c</sup> $\eta^2$ effect size benchmarks: negligible ( $\eta^2 < 0.01$ ), small ( $0.01 \leq \eta^2 < 0.06$ ), medium ( $0.06 \leq \eta^2 < 0.14$ ), and large ( $\eta^2 \geq 0.14$ ) partial effect sizes.

### 3.3. Baseline ODI predicting follow-up mood symptoms

Greater baseline ODI of the uncinate fasciculus was significantly associated with higher mania symptoms at 6-month follow-up (Table 3). Trend-level associations were also observed between higher ODI in the cingulum bundle and forceps minor and subsequent mania symptoms. No significant associations were found between baseline ODI and depressive symptoms (Table 3).

**Table 3.** Relationship between ODI of white matter tracts involved in emotion regulation and mood symptoms 6 months later.

| Independent Variable | Outcome | $\beta$ | P<sup>a</sup> | Q<sup>a,b</sup> | $\eta^2$<sup>c</sup> |
| --- | --- | --- | --- | --- | --- |
| Cingulum Bundle ODI | Mania symptoms (YMRS) | 0.21 | *0.060* | 0.120 | 0.06 |
|  | Depressive symptoms (HRSD) | -0.09 | 0.340 | 0.340 | 0.03 |
| Forceps Minor ODI | Mania symptoms (YMRS) | 0.23 | **0.043** | *0.086* | 0.06 |
|  | Depressive symptoms (HRSD) | -0.06 | 0.572 | 0.572 | 0.01 |
| Uncinate Fasciculus ODI | Mania symptoms (YMRS) | 0.25 | **0.023** | **0.046** | 0.06 |
|  | Depressive symptoms (HRSD) | <0.01 | 0.995 | 0.995 | <0.01 |

Abbreviations: ODI – Orientation Dispersion Index; YMRS – Young Mania Rating Scale; HRSD – Hamilton Rating Scale for Depression.
<sup>a</sup> P-values $\leq 0.05$ are reported in bold characters; P-values between 0.05 and 0.10 are reported in italics.
<sup>b</sup> Q-values indicate P-values after False Discovery Rate (FDR) correction for multiple comparisons. FDR correction was applied separately for each independent variable across all outcomes it was tested on.
<sup>c</sup> $\eta^2$ effect size benchmarks: negligible ( $\eta^2 < 0.01$ ), small ( $0.01 \leq \eta^2 < 0.06$ ), medium ( $0.06 \leq \eta^2 < 0.14$ ), and large ( $\eta^2 \geq 0.14$ ) partial effect sizes.

### 3.4. Mediation analyses

ODI of the uncinate fasciculus fully mediated the association between CFI at baseline and mania symptoms six months later (β=-0.10,95%CI[-0.14,-0.02]). Specifically, lower CFI was associated with higher ODI, particularly in those with greater mania vulnerability, which in turn was associated with more mania symptoms six months later (Figure 2).

**Figure 2.**
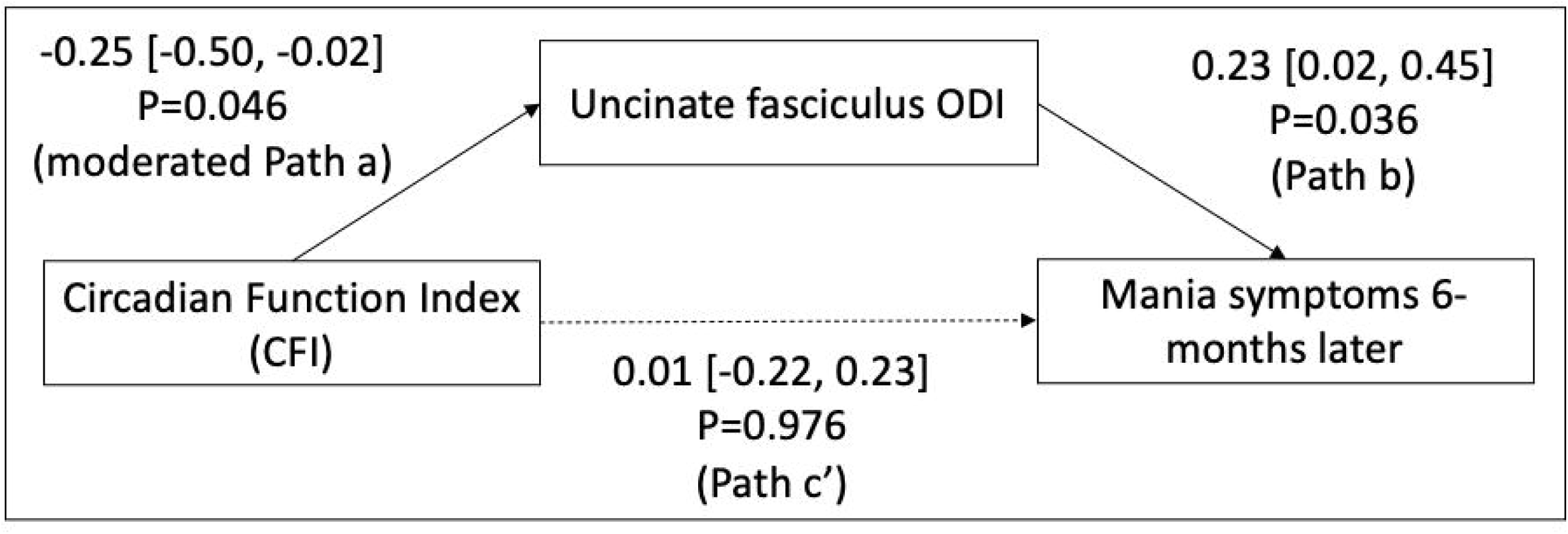
Mediation findings linking rest-activity rhythms instability, brain white matter, and mania symptoms six months later. Mediation model linking baseline Circadian Function Index (CFI; independent variable), baseline Uncinate Fasciculus Orientation Dispersion Index (ODI; mediator), and mania symptoms 6 months later (outcome). Arrows indicate direction of effects; dashed lines represent nonsignificant associations. Regression coefficients, confidence intervals, and p-values are shown. Lower CFI predicted higher Uncinate Fasciculus ODI (Path a), moderated by mania vulnerability (MOODS-SRL). Higher ODI predicted more mania symptoms at follow-up (Path b), while the direct effect of CFI on mania symptoms (Path c’) was nonsignificant. The indirect effect of CFI on mania symptoms via ODI was significant, β = -0.10, 95% CI [-0.14, -0.02].

### 3.5. Exploratory analyses

#### CFI components

Follow-up analyses suggested that greater rhythm fragmentation (IV) was linked to higher ODI in both the cingulum bundle and uncinate fasciculus, while lower rhythm stability (IS) was associated with higher ODI only in the uncinate fasciculus, with no effects of rhythm amplitude (RA). See Supplemental Materials for more information.

#### Tract-profile analyses

Tract-profile models indicated that the moderated effect of CFI on ODI in the cingulum bundle was strongest in a specific portion of the tract, namely the posterior termination (Figure 1-Panel B; Supplemental Table 1). In addition, the moderated effect of CFI on ODI in the uncinate fasciculus was most pronounced in the prefrontal termination of the tract (Figure 1-Panel E; Supplemental Table 1).

#### Other sleep and rest-activity rhythms metrics

Exploratory analyses indicated that mania vulnerability moderated the link between later lights-off time and higher ODI in the uncinate fasciculus, but ODI did not mediate the association between lights-out time and mood symptoms. A similar moderation trend for later sleep onset did not survive correction, and no other sleep or rest-activity metrics were significant (Supplemental Table 2). See Supplemental Materials for more information.

#### Other diffusion metrics and white matter tracts

There were no significant associations involving NDI or FA (Supplemental Table 3). In addition, mania vulnerability did not moderate the relationships between sleep or rest-activity rhythms instability and ODI metrics of other white matter tracts (Supplemental Table 4).

## 4. DISCUSSION

Consistent with our first hypothesis, rest-activity rhythm instability (lower CFI) was associated with reduced white matter coherence (higher ODI) in the cingulum bundle and uncinate fasciculus, especially in individuals with higher mania vulnerability (MOODS-SRL). Supporting our second hypothesis, higher ODI in the uncinate fasciculus predicted greater mania symptoms (YMRS) six months later, with mediation analyses indicating that this tract fully mediated the CFI-mania link. Importantly, we found no associations involving sleep duration or timing instability, and CFI was specifically linked to mania but not depression models, underscoring a selective connection between rest-activity rhythm instability and mania symptoms. Together with prior work(Lima Santos et al., 2025), our results extend this literature by providing initial evidence that instability in rest-activity rhythms may contribute to BD vulnerability through its effects on white matter organization within emotion regulation circuits.

Notably, our moderation pattern suggests that rest-activity rhythm instability is more strongly associated with white matter microstructure in individuals with higher mania vulnerability, indicating a potential heightened susceptibility in those at greater BD risk. One plausible pathway comes from animal work showing that circadian instability can alter oligodendrocyte activity and myelination, ultimately compromising axonal organization(Colwell & Ghiani, 2020; Tang et al., 2025; Zuo et al., 2024). However, given the cross-sectional nature of these moderation models, we cannot exclude the possibility that the relationship occurs in the opposite direction, with higher ODI contributing to greater CFI. Nonetheless, little is known about the factors that could make a subset of individuals particularly vulnerable to these associations. One possibility is that BD has been associated with alterations in hormonal function and inflammation(Lyu et al., 2023; Muneer, 2015), which may increase sensitivity of white matter to fluctuations in daily rhythms and vice versa. Altogether, these findings underscore the dynamic link between circadian rhythm stability and BD risk factors in relation to white matter organization and highlight the importance of ongoing efforts to stabilize modifiable factors, such as rest-activity rhythms. Future longitudinal studies integrating neuroimaging with detailed circadian and sleep assessments, as well as direct measurements of possible biological factors (e.g., hormonal and inflammatory markers), are needed to clarify the temporal and mechanistic nature of these associations.

Our finding that lower white matter coherence (greater ODI) in the uncinate fasciculus predicted greater mania symptoms 6-months later suggests that this heightened microstructural sensitivity could accelerate the progression from subthreshold to syndromal mood symptoms. Given that circadian instability is common, especially during adolescence(Kolip et al., 2022; Wallace et al., 2023), such disruptions may heighten vulnerability even amid normative developmental changes in rhythm regulation among individuals at risk for BD. Furthermore, this also underscores the clinical relevance of circadian stability. Stabilizing rest-activity rhythms may help preserve white matter integrity, which, in the context of BD risk, could mitigate microstructural alterations in emotion regulation pathways that contribute to future mood dysregulation. Given that white matter abnormalities often precede the onset of BD(Drevets et al., 2008; Goldstein & Walker, 2014; Harrison et al., 2018; Ng et al., 2009; Tkachev et al., 2003), interventions targeting behavioral rhythm stability may hold promise for prevention or early intervention. This is supported by previous studies showing that, white matter structure appears sensitive to therapeutic modulation, particularly to mood-stabilizing medications such as lithium(Espanhol & Vieira-Coelho, 2022), which underscores its plasticity and potential role as a treatment target. Together, these findings support the notion that rhythm-stabilizing strategies, including behavioral and pharmacological approaches, may protect both brain and emotional health in individuals at risk for BD.

More broadly, our findings advance our understanding of the links between rest-activity rhythm and brain white matter. Notably, microstructural alterations were more strongly linked to rhythm fragmentation (IV) and reduced stability (IS) than to amplitude (RA), suggesting that day-to-day irregularity, rather than overall activity level, may be the key circadian feature affecting white matter. While sleep is known to support neuronal maintenance, glial function, and myelin repair(Bellesi, 2015; de Vivo & Bellesi, 2019), few studies have focused on circadian rhythms as a distinct driver of white matter integrity. Prior work shows that greater instability in RA, IS, and IV is associated with lower FA and that white matter mediates links between circadian instability and cognition in older adults(Callow et al., 2025; McMahon et al., 2021). Our study is the first to link circadian instability to NODDI metrics, offering several insights: (1) it provides a mechanistic link between behavioral rhythms and specific aspects of microstructure, particularly white matter coherence, with NODDI potentially more sensitive than FA in capturing microstructural changes related to circadian rhythm; (2) associations were confined to emotion regulation tracts, suggesting selective vulnerability of affective networks; and (3) circadian instability may influence emotional and cognitive outcomes across the lifespan via effects on white matter.

Although these findings offer valuable insights, some limitations should be noted. First, the 6-month follow-up was relatively short, limiting assessment of long-term mood outcomes and bidirectional relationships. Longer follow-ups are needed to determine whether observed associations persist. Second, because participants did not have clinical mood disorders, these results may not generalize to individuals with diagnosed bipolar disorder. Finally, although our mediation model suggested that ODI in the uncinate fasciculus fully mediated the association between CFI and follow-up mania symptoms, this result should be interpreted with caution. Notably, the direct path from CFI to later mania symptoms was not significant in the absence of the mediator. While this pattern is not uncommon in mechanistic models in which a behavioral predictor influences outcomes primarily through a more proximal biological process, it limits the strength of inferences that can be drawn about a direct relationship. Larger samples and longer follow-up will help clarify whether CFI directly predicts later mania or acts mainly through white matter changes.

Altogether, our findings provide converging evidence that circadian rhythm instability plays a critical role in shaping white matter microstructure in individuals at heightened risk for BD. This microstructural sensitivity, in turn, predicted future increases in mania symptoms, suggesting a potential pathway through which circadian instability may accelerate progression from subthreshold to syndromal mood dysregulation. Clinically, these results underscore the relevance of circadian stability as a potential target for preventive or early interventions, including behavioral and pharmacological strategies aimed at preserving white matter integrity in emotion regulation circuits. More broadly, our study extends prior work by demonstrating that circadian instability is linked to fine-grained NODDI measures, offering insight into the microstructural pathways that may connect rhythm dysregulation to emotional and cognitive outcomes. Future longitudinal studies incorporating detailed circadian, sleep, and biological assessments are needed to clarify the mechanisms and temporal dynamics of these relationships, which may ultimately inform strategies to protect brain health and mitigate risk for BD.

## Data Availability

Data may be made available from the corresponding author on reasonable request.

