## Supplemental Materials for "White matter microstructure mediates the link between rest-activity rhythm instability and mania symptoms in individuals at risk for bipolar disorder"

**SUPPLEMENTAL METHODS.**

***Participants***. Exclusion criteria included 1) Unable to read and write in English; 2) IQ < 70; 3) Left or mixed handedness; 4) Current/lifetime bipolar spectrum disorder or psychosis spectrum disorder; 5) Mood stabilizer or antipsychotic medication use and/or changes to psychotropic medication in the past 2 months; 6) Substance or alcohol use disorder and/or illicit substance use [except cannabis] in the past 3 months; 7) History of head trauma, systemic medical illness, or ophthalmological or neurological conditions; 8) Organic sleep disorders, including sleep apnea, restless legs syndrome, or narcolepsy; 9) Current shift work; 10) MRI contraindication (e.g., metals in the body, recent tattoo, claustrophobia); 11) Pregnant or breastfeeding women; 12) Extreme sleep schedules (for example, bedtime > 3am and waketime > 11am, or bedtime < 9pm and waketime < 5am); 13) Color blindness, red-green color deficiency, or other rarer forms of abnormal color vision if they have potential to impact study outcome measures (<9 on the Ishihara's Test of Color Blindness); 14) A family member previously enrolled in the study.

***Neuroimaging.*** Additional white matter tracts included: Anterior thalamic radiation, Arcuate fasciculus, Cortical spinal tract, Inferior longitudinal fasciculus, Inferior occipital-frontal tract, Striato-fronto-orbital tract, Superior longitudinal fasciculus, Superior thalamic radiation, Thalamo-parietal tract, Thalamo-premotor, and Thalamo-occipital tract.

**SUPPLEMENTAL RESULTS.**

***Exploratory analyses - CFI components.*** Follow-up analyses examining individual CFI components indicated that greater IV (i.e., more fragmented rest–activity rhythms) was associated with greater ODI in both the cingulum bundle (β=0.25,P=0.012,Q=0.012) and the uncinate fasciculus (β=0.26,P=0.006,Q=0.012). In contrast, lower IS (i.e., lower rest–activity rhythm stability) was associated with greater ODI exclusively in the uncinate fasciculus (β=-0.23,P=0.024,Q=0.048), with no corresponding association in the cingulum bundle (β=-0.17,P=0.113,Q=0.113). There were no significant associations involving RA in either tract (cingulum bundle: β=-0.08,P=0.509, Q=0.572; uncinate fasciculus: β=-0.06,P=0.572,Q=0.572).

***Exploratory analyses - Other sleep and rest-activity rhythms*** ***metrics.*** Exploratory models revealed a significant moderation effect of mania vulnerability on the association between later lights-off time and higher ODI in the uncinate fasciculus (Supplemental Table 2). However, ODI of the uncinate fasciculus did not mediate the association between lights-out time and depressive (β=-0.01,95%CI[-0.07,0.08]) or mania symptoms at follow-up (β=0.07,95%CI[-0.01,0.22]). Additionally, mania vulnerability moderated the association between later sleep onset and higher ODI in the uncinate fasciculus, although this association did not survive FDR correction (Supplemental Table 2). No significant associations were found for other sleep and rest-activity rhythms metrics (Supplemental Table 2).

**SUPPLEMENTAL TABLES.**

**Supplemental Table 1. Tract-profile analyses to identify the strongest association between CFI and ODI of white matter tracts involved in emotion regulation.**

| White matter tract | Nodes | **β** | **P ^a^** | **Q ^a,b^** |
| --- | --- | --- | --- | --- |
| Cingulum Bundle ^c^ | Node 1 | -0.17 | *0.067* | *0.089* |
|  | Node 2 | -0.16 | *0.071* | *0.089* |
|  | Node 3 | -0.19 | **0.033** | *0.083* |
|  | Node 4 | -0.15 | *0.092* | *0.092* |
|  | Node 5 | -0.24 | **0.009** | **0.045** |
| Uncinate Fasciculus ^d^ | Node 1 | -0.09 | 0.298 | 0.298 |
|  | Node 2 | -0.18 | *0.044* | *0.078* |
|  | Node 3 | -0.18 | *0.047* | *0.078* |
|  | Node 4 | -0.13 | 0.150 | 0.188 |
|  | Node 5 | -0.23 | **0.008** | **0.040** |

^a^ P-values ≤ 0.05 are reported in bold characters; P-values between 0.05 and 0.10 are reported in italics.

^b^ Q-values indicate P-values after False Discovery Rate (FDR) correction for multiple comparisons. FDR correction was applied separately for each white matter tract.

^c^ In the cingulum bundle, nodes 1 to 5 are ordered from anterior to posterior.

^d^ In the uncinate fasciculus, nodes 1 to 5 are ordered from temporal to prefrontal.

**Supplemental Table 2. Moderated association between the average of other sleep variables and ODI metrics of white matter tracts involved in emotion regulation.**

| **Independent Variable** | **Outcome** | **β** | **P ^a^** | **Q ^a,b^** | **η2 ^c^** |
| --- | --- | --- | --- | --- | --- |
| Sleep duration | Cingulum Bundle ODI | 0.01 | 0.903 | 0.903 | <0.01 |
|  | Forceps Minor ODI | 0.09 | 0.405 | 0.839 | 0.01 |
|  | Uncinate Fasciculus ODI | -0.06 | 0.559 | 0.839 | <0.01 |
| Sleep onset | Cingulum Bundle ODI | -0.01 | 0.924 | 0.924 | <0.01 |
|  | Forceps Minor ODI | 0.07 | 0.510 | 0.765 | <0.01 |
|  | Uncinate Fasciculus ODI | 0.21 | **0.038** | 0.114 | 0.04 |
| Lights out | Cingulum Bundle ODI | 0.01 | 0.925 | 0.925 | <0.01 |
|  | Forceps Minor ODI | 0.08 | 0.414 | 0.621 | 0.01 |
|  | Uncinate Fasciculus ODI | 0.25 | **0.009** | **0.027** | 0.07 |
| Get up time | Cingulum Bundle ODI | 0.00 | 0.978 | 0.978 | <0.01 |
|  | Forceps Minor ODI | 0.11 | 0.267 | 0.400 | 0.01 |
|  | Uncinate Fasciculus ODI | 0.13 | 0.184 | 0.400 | 0.02 |

Abbreviations: ODI – Orientation Dispersion Index.

^a^ P-values ≤ 0.05 are reported in bold characters.

^b^ Q-values indicate P-values after False Discovery Rate (FDR) correction for multiple comparisons. FDR correction was applied separately for each independent variable across all outcomes it was tested on.

^c^ η2 effect size benchmarks: negligible (η2 < 0.01), small (0.01 ≤ η2 < 0.06), medium (0.06 ≤ η2 < 0.14), and large (η2 ≥ 0.14) partial effect sizes.

**Supplemental Table 3. Moderated association between sleep or rest-activity rhythms instability and NDI/FA of white matter tracts involved in emotion regulation.**

| **NDI** | | | | | |
| --- | --- | --- | --- | --- | --- |
| **Independent Variable** | **Outcome** | **β** | **P** | **Q ^a^** | **η2 ^a^** |
| Sleep duration  variability | Cingulum Bundle ODI | -0.15 | 0.176 | 0.257 | 0.02 |
|  | Forceps Minor ODI | -0.15 | 0.176 | 0.257 | 0.02 |
|  | Uncinate Fasciculus ODI | -0.12 | 0.257 | 0.257 | 0.01 |
| Sleep onset  variability | Cingulum Bundle ODI | -0.13 | 0.144 | 0.216 | 0.02 |
|  | Forceps Minor ODI | -0.14 | 0.119 | 0.216 | 0.03 |
|  | Uncinate Fasciculus ODI | -0.05 | 0.560 | 0.560 | <0.01 |
| CFI | Cingulum Bundle ODI | 0.03 | 0.791 | 0.791 | <0.01 |
|  | Forceps Minor ODI | -0.04 | 0.694 | 0.791 | <0.01 |
|  | Uncinate Fasciculus ODI | -0.11 | 0.234 | 0.702 | 0.02 |
| **FA** | | | | | |
| **Independent Variable** | **Outcome** | **β** | **P** | **Q ^a^** | **η2 ^b^** |
| Sleep duration  variability | Cingulum Bundle ODI | -0.14 | 0.223 | 0.410 | 0.02 |
|  | Forceps Minor ODI | -0.01 | 0.911 | 0.911 | <0.01 |
|  | Uncinate Fasciculus ODI | -0.12 | 0.273 | 0.410 | 0.01 |
| Sleep onset  variability | Cingulum Bundle ODI | -0.12 | 0.196 | 0.588 | 0.02 |
|  | Forceps Minor ODI | 0.00 | 0.971 | 0.971 | <0.01 |
|  | Uncinate Fasciculus ODI | -0.02 | 0.785 | 0.971 | <0.01 |
| CFI | Cingulum Bundle ODI | 0.17 | 0.084 | 0.209 | 0.03 |
|  | Forceps Minor ODI | 0.12 | 0.209 | 0.209 | 0.02 |
|  | Uncinate Fasciculus ODI | 0.13 | 0.148 | 0.209 | 0.02 |

Abbreviations: NDI – Neurite Density Index; FA – Fractional anisotropy; CFI – Circadian Function Index.

^a^ Q-values indicate P-values after False Discovery Rate (FDR) correction for multiple comparisons. FDR correction was applied separately for each independent variable across all outcomes it was tested on.

^b^ η2 effect size benchmarks: negligible (η2 < 0.01), small (0.01 ≤ η2 < 0.06), medium (0.06 ≤ η2 < 0.14), and large (η2 ≥ 0.14) partial effect sizes.

**Supplemental Table 4. Moderated association between the sleep or rest-activity rhythms** **instability and ODI metrics of other white matter tracts.**

| **Independent Variable** | **Outcome** | **β** | **P ^a^** | **Q ^b^** |
| --- | --- | --- | --- | --- |
| Sleep duration variability | Anterior thalamic radiation ODI | 0.09 | 0.371 | 0.794 |
|  | Arcuate fasciculus ODI | -0.07 | 0.505 | 0.794 |
|  | Cortical spinal tract ODI | -0.12 | 0.301 | 0.794 |
|  | Inferior longitudinal fasciculus ODI | -0.08 | 0.467 | 0.794 |
|  | Inferior occipital-frontal tract ODI | 0.01 | 0.953 | 0.953 |
|  | Striato-fronto-orbital tract ODI | 0.02 | 0.816 | 0.898 |
|  | Superior longitudinal fasciculus ODI | -0.03 | 0.776 | 0.898 |
|  | Superior thalamic radiation ODI | -0.19 | *0.092* | 0.794 |
|  | Thalamo-parietal tract ODI | -0.11 | 0.297 | 0.794 |
|  | Thalamo-premotor ODI | -0.09 | 0.435 | 0.794 |
|  | Thalamo - occipital tract ODI | 0.04 | 0.680 | 0.898 |
| Sleep onset variability | Anterior thalamic radiation ODI | 0.03 | 0.739 | 0.813 |
|  | Arcuate fasciculus ODI | -0.09 | 0.360 | 0.731 |
|  | Cortical spinal tract ODI | -0.12 | 0.202 | 0.731 |
|  | Inferior longitudinal fasciculus ODI | -0.12 | 0.212 | 0.731 |
|  | Inferior occipital-frontal tract ODI | -0.02 | 0.850 | 0.850 |
|  | Striato-fronto-orbital tract ODI | -0.05 | 0.606 | 0.741 |
|  | Superior longitudinal fasciculus ODI | -0.05 | 0.580 | 0.741 |
|  | Superior thalamic radiation ODI | -0.14 | 0.155 | 0.731 |
|  | Thalamo-parietal tract ODI | -0.10 | 0.297 | 0.731 |
|  | Thalamo-premotor ODI | -0.07 | 0.465 | 0.731 |
|  | Thalamo-occipital tract ODI | -0.07 | 0.412 | 0.731 |
| CFI | Anterior thalamic radiation ODI | -0.16 | *0.065* | 0.363 |
|  | Arcuate fasciculus ODI | 0.03 | 0.725 | 0.830 |
|  | Cortical spinal tract ODI | 0.02 | 0.817 | 0.830 |
|  | Inferior longitudinal fasciculus ODI | 0.06 | 0.505 | 0.830 |
|  | Inferior occipital-frontal tract ODI | -0.12 | 0.171 | 0.586 |
|  | Striato-fronto-orbital tract ODI | -0.16 | *0.066* | 0.363 |
|  | Superior longitudinal fasciculus ODI | -0.05 | 0.629 | 0.830 |
|  | Superior thalamic radiation ODI | -0.09 | 0.370 | 0.814 |
|  | Thalamo-parietal tract ODI | -0.02 | 0.830 | 0.830 |
|  | Thalamo-premotor ODI | -0.12 | 0.213 | 0.586 |
|  | Thalamo - occipital tract ODI | -0.03 | 0.785 | 0.830 |

Abbreviations: ODI – Orientation Dispersion Index; CFI – Circadian Function Index.

^a^ P-values between 0.05 and 0.10 are reported in italics.

^b^ Q-values indicate P-values after False Discovery Rate (FDR) correction for multiple comparisons. FDR correction was applied separately for each independent variable across all outcomes it was tested on.

**SUPPLEMENTAL FIGURES.**

**Supplemental Figure 1. Distribution of MOODS-SRL scores.**

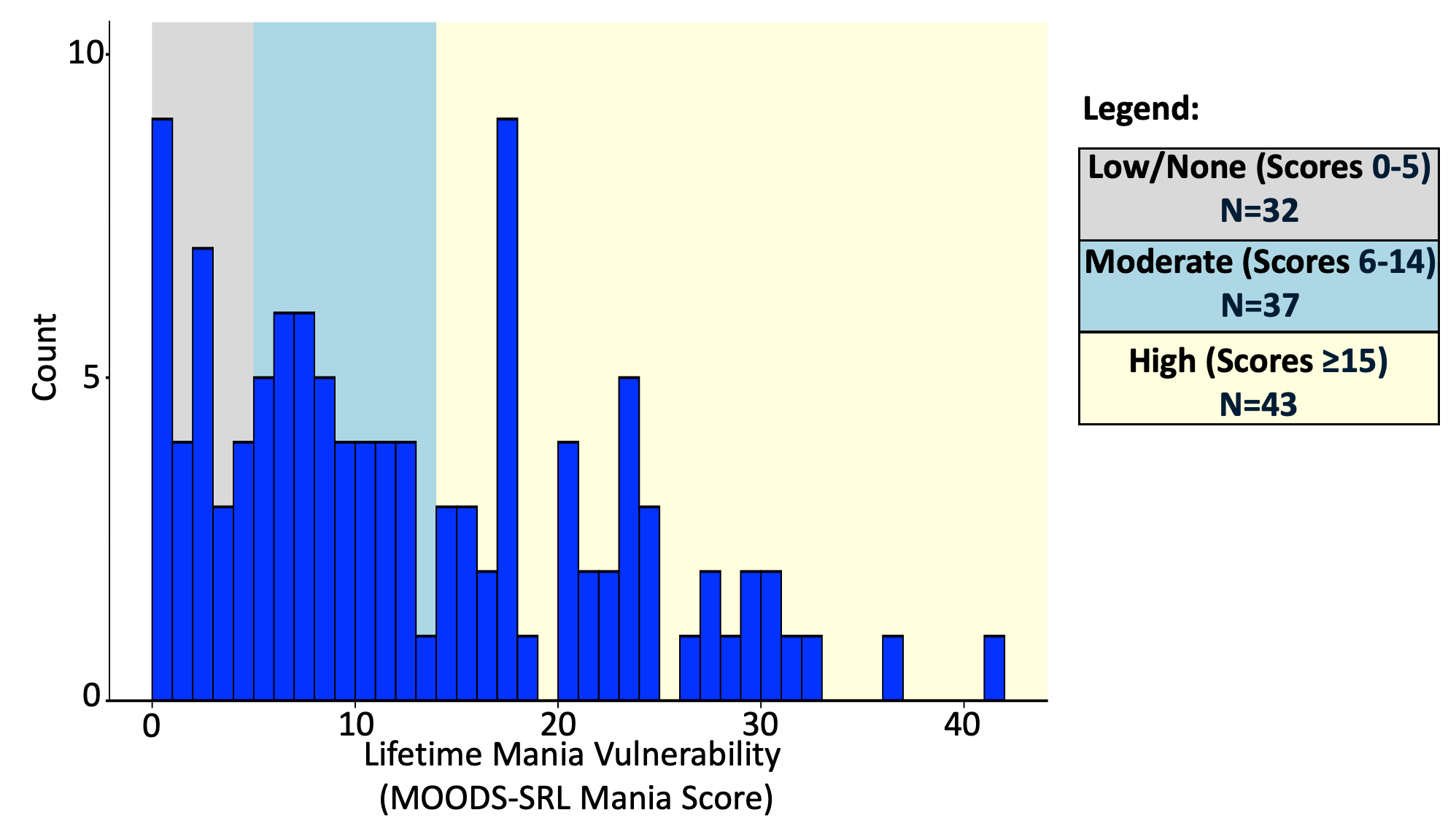

**Supplemental Figure 1 legend.** Distribution of MOODS-SRL mania scores at baseline. Bars represent the frequency of scores. Shaded bands indicate severity ranges: none/low (0–5; gray), moderate (6–14; blue), and high (≥15; yellow).
